# Effectiveness of Bivalent Boosters Over Nine and Half Months in Nebraska

**DOI:** 10.1101/2023.12.03.23299338

**Authors:** Sai Paritala, Yangjianchen Xu, Yi Du, Matthew Donahue, Patrick Maloney, Dan-Yu Lin

## Abstract

On September 1, 2022, Moderna and Pfizer–BioNTech bivalent vaccines replaced existing monovalent vaccines as booster doses against severe acute respiratory syndrome coronavirus 2 (SARS-CoV-2) for persons aged 12 and older in the United States. We assessed the effectiveness of these bivalent boosters against Omicron infection and severe outcomes (COVID-19 hospitalization and death) over a 9.5-month period using line-level data from the state of Nebraska. We found that the relative effectiveness of bivalent boosters, compared with one fewer vaccine dose, against Omicron infection and subsequent death was 39.0% (95% confidence interval [CI], 35.7 to 42.2) and 70.0% (95% CI, 38.8 to 85.3), respectively, at four weeks post administration and gradually waned afterward. Vaccine effectiveness was broadly similar against different Omicron subvariants.

## Introduction

On August 31, 2022, the Food and Drug Administration authorized the Moderna and Pfizer– BioNTech bivalent COVID-19 vaccines, each containing equal amounts of mRNA encoding the spike protein from the ancestral strain and the spike protein from the BA.4 and BA.5 strains of the B.1.1.529 (Omicron) variant, for emergency use as a single booster dose at least 2 months after primary or booster vaccination.^1^ On September 1, 2022, bivalent mRNA vaccines replaced monovalent vaccines as booster doses for persons 12 years of age or older in the United States. We conducted an investigation into the effectiveness of these two bivalent boosters during the first 9.5 months of deployment (September 1, 2022 to June 15, 2023) using line-level data on 754,758 Nebraska residents who were eligible to receive bivalent boosters. During this 9.5-month period, the Omicron variant was predominant, with the circulating strains evolving from BA.4/BA.5 to BQ.1/BQ.1.1 and finally to XBB/XBB.1.5.

## Methods

### Study Design

We obtained line-level data on COVID-19 vaccination from December 11, 2020 to June 15, 2023 and on COVID-19 infection, hospitalization, and death from March 3, 2020 to June 15, 2023. The data were obtained through linkage of the Nebraska Electronic Disease Surveillance System, Nebraska State Immunization Information System, and Nebraska Hospital Discharge Data.

Records were probabilistically linked, where a probabilistic score was assigned to each record to determine the quality of the match. Scores were developed using unique data elements, including first name, last name, middle name, date of birth, gender, and residential zip code. Records with high match scores were linked and included in the final dataset.

Our primary objective was to examine the association between bivalent booster vaccination and Omicron infection, hospitalization, and death. We focused on the period between September 1, 2022, and June 15, 2023. COVID-19 infection and vaccination before September 1, 2022, were included as covariates. Individuals included in this study were aged 12 or older and had completed a primary vaccination series before September 1, 2022.

COVID-19 infections were captured via mandated reports from Nebraska Electronic Laboratory Reporting records statewide. At-home tests were not included. COVID-19 hospitalizations were captured via Nebraska Hospital Discharge data and COVID-19 case investigation. COVID-19 deaths were initially extracted from COVID-19 case investigations and then validated via death certificates and electronic laboratory reporting records review and/or contacting physicians, coroners, and/or patient relatives.

### Statistical Analysis

We considered four endpoints: (1) recurrent infection times; (2) time to hospitalization; (3) time to hospitalization or death, whichever occurs first; and (4) time to death. For endpoints (2) to (4), we fit the Cox regression model in which the log hazard ratio for each additional booster dose that was received (i.e., first booster vs. primary vaccination, second booster vs. first booster, or third booster vs. second booster) is a continuous B-spline function of the time elapsed since receipt of a booster dose.^2-5^ To ascertain the ramping and waning patterns, we used a piecewise linear function with change points at 2 and 4 weeks (i.e., 14 and 28 days) after receipt of a booster dose. To reduce confounding bias due to time trends in disease incidence, we measured the event time for each person from the start of the study period. This allowed us to compare the risks of disease for boosted and non-boosted persons at the same point in time on the calendar. To further reduce confounding bias, we included vaccine manufacturer and date of previous vaccination (and the product between these two variables), previous infection status (yes or no), and demographic factors (i.e., sex, age group, race/ethnicity, and socioeconomic status) as covariates. We also included the vaccination status at the start of the study period (receipt of only the primary vaccine series, receipt of only the first booster dose, or receipt of two booster doses) as two indicator covariates, allowing individuals with different baseline vaccination statuses to have different disease risks.

When analyzing the second endpoint, we censored hospitalization at time of death, such that the Cox regression analysis pertained to cause-specific hazard function, rather than the usual hazard function.^6^ For the first endpoint, we used the proportional rates model for recurrent events instead of the Cox proportional hazards model for a single event.^7^

We first analyzed the data for the entire period of bivalent boosters, i.e., September 1, 2022 to June 15, 2023. We then analyzed the data separately for individuals who received bivalent boosters before November 1, 2022 (when BA.4/BA.5 were predominant) and after November 1, 2022 (when BQ.1/BQ.1.1 became more prevalent and then were gradually replaced by XBB/XBB.1.5).

The effect of an additional booster dose was characterized by the hazard ratio function, HR(t), in the Cox proportional hazards model or the rate ratio function, RR(t), in the proportional rates model. The relative vaccine effectiveness of a bivalent booster (compared with one fewer vaccine dose) at time t, VE(t), was defined to be 100x[1 − HR(t)]% or 100x[1 − RR(t)]%.^8^ Maximum partial likelihood is used to estimate HR(t), RR(t), and VE(t), and 95% confidence intervals (CI) were constructed.

## Results

From September 1, 2022 to June 15, 2023, a lot of 754,758 individuals in the state of Nebraska were eligible for bivalent boosters, and 215,567 (28.6%) received them; 5,495 (22.6%) of the 24,297 SARS-CoV-2 infections, 371 (26.9%) of the 1,380 Covid-19 related hospitalizations, and 59 (26.1%) of the 226 Covid-19 related deaths occurred after receipt of the bivalent booster (Table 1).

**Table 1.**
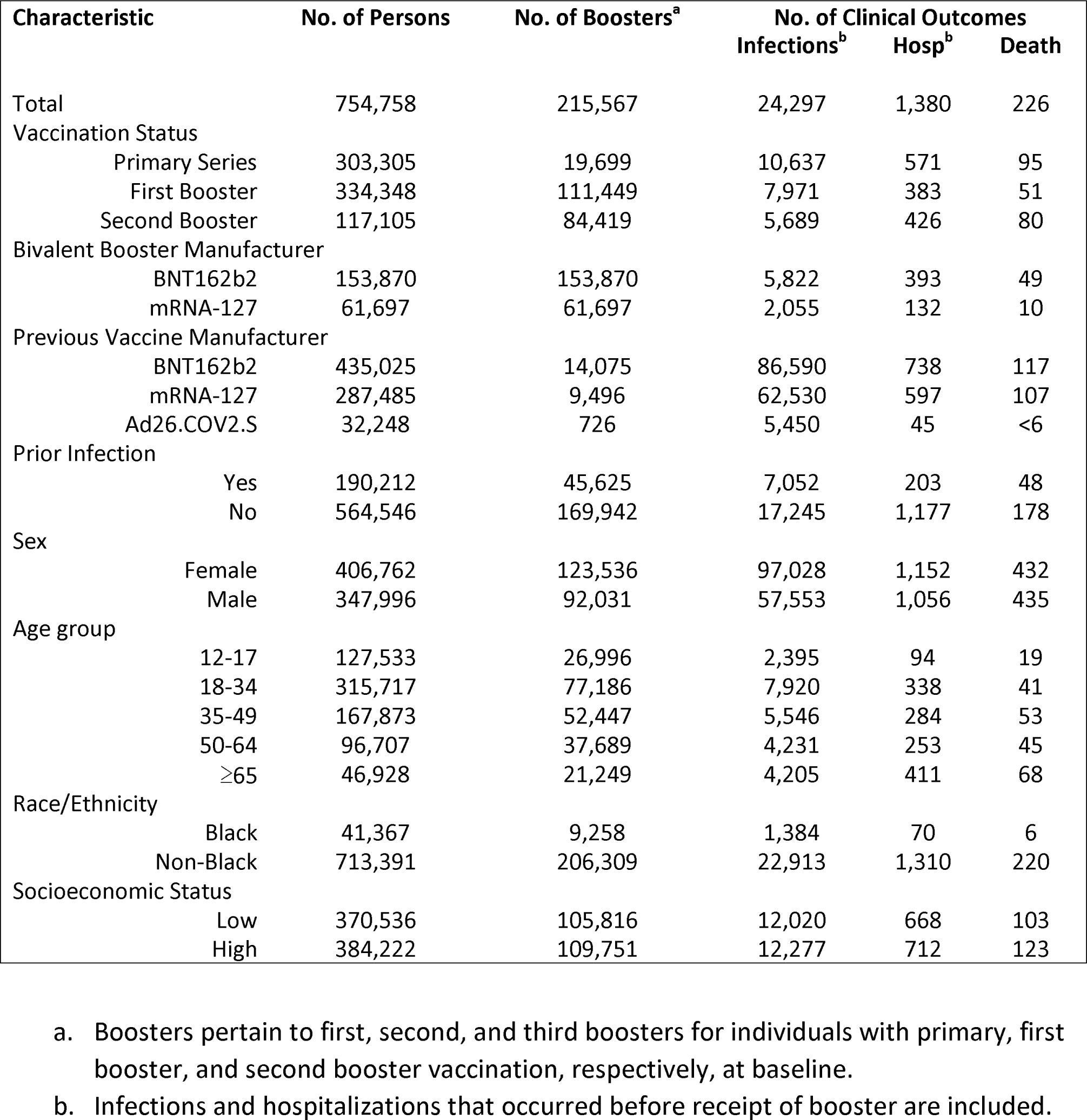
Demographic and Clinical Characteristics of Analysis Population.

The estimation results for the relative effectiveness of bivalent boosters (compared with one fewer vaccine dose) during the entire study period are shown in Figure 1 (left column). Relative effectiveness against infection reached a level of 39.0% (95% CI, 35.7 to 42.2) 4 weeks post injection and decreased to 28.9% (95% CI, 25.9 to 31.9), 17.2% (95% CI, 14.2 to 20.1) and 3.6% (95% CI, 0.2 to 6.9) after 8, 12 and 16 weeks, respectively (Fig. 1 A). Relative effectiveness against hospitalization reached 43.6% (95% CI, 26.0 to 57.1) after 4 weeks and decreased to 38.8% (95% CI, 26.4 to 49.2), 33.6% (95% CI, 19.4 to 45.4), 28.0% (95% CI, 3.6 to 46.2) and 21.8% (95% CI, 0.0 to 48.6) after 8, 12, 16, and 20 weeks, respectively (Fig. 1 B). Relative effectiveness against hospitalization or death reached a level of 46.8% (95% CI, 30.9 to 59.0) after 4 weeks and decreased to 40.4% (95% CI, 28.9 to 50.1), 33.3% (95% CI, 19.9 to 44.5), 25.4% (95% CI, 1.9 to 43.2), and 16.5% (95% CI, 0.0 to 43.7) after 8 weeks, 12 weeks, 16 weeks, and 20 weeks, respectively (Fig. 1 E). Relative effectiveness against death reached a level of 70.0% (95% CI, 38.8 to 85.3) after 4 weeks and decreased to 58.0% (95% CI, 32.2 to 73.9), 50.3% (95% CI, 24.0 to 67.5), and 17.6% (95% CI, 0.0 to 56.1) after 8 weeks, 12 weeks and 16 weeks, respectively (Fig. 1 G).

**Figure 1.**
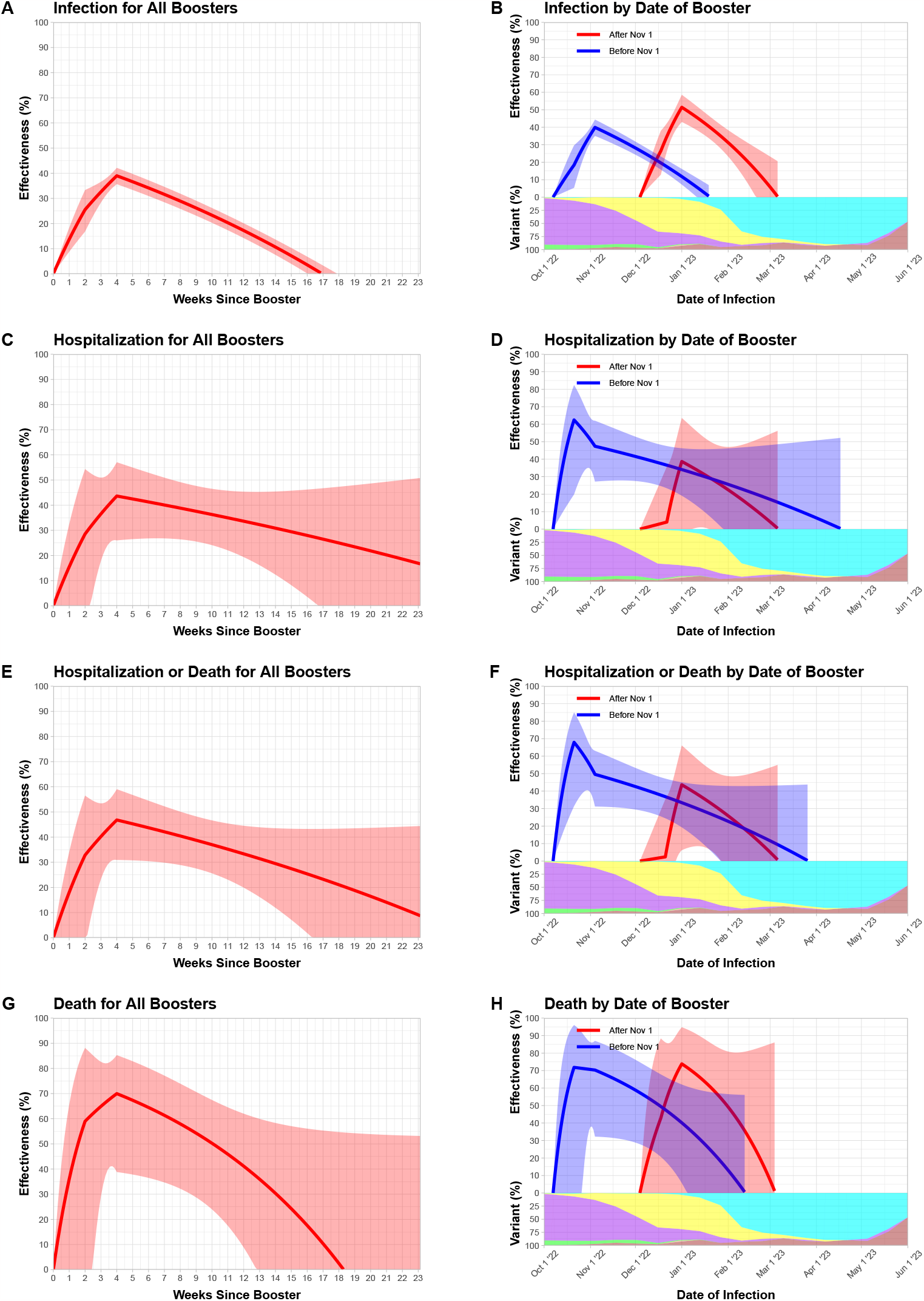
Relative effectiveness of A Single Bivalent Booster Dose as a Function of Time Elapsed Since Receipt of the Booster. The first, second, third, and fourth rows pertain the endpoints of infection, hospitalization, hospitalization or death, and death, respectively. The left column pertains to the analysis of all bivalent booster doses, and the right column pertains to the stratified analysis by booster cohort (i.e., receipt date of the booster dose). The log hazard ratio or rate ratio for a bivalent booster dose (compared with one less dose) is approximated by a continuous, piecewise linear function with change points at 2 and 4 weeks after receipt of the booster. The solid curves show the estimates of (relative) booster effectiveness. The shaded bands indicate 95% confidence intervals. In (B), (D), (F), and (H), each curve starts at the median receipt date of the booster dose for individuals in that cohort, and the proportions of BA.4, BA.5, BQ.1/BQ.1.1, XBB/XBB.1.5, and other strains are indicated by the green, purple, yellow, cyan, and brown areas, respectively.

The results for the two sub-periods, i.e., before November 1, 2022 versus after November 1, 2022 are shown in Figure 1 (right column). Before November 1, 2022, relative effectiveness against infection was 40.0% (95% CI, 35.1 to 44.4), 27.7% (95% CI, 23.9 to 31.3) and 13.0% (95% CI, 8.5 to 17.2) after 4 weeks, 8 weeks and 12 weeks, respectively. After November 1, 2022, relative effectiveness against infection was 51.4% (95% CI, 43.0 to 58.6), 33.5% (95% CI, 26.1 to 40.2) and 9.0% (95% CI, 0.0 to 24.8) after 4 weeks, 8 weeks and 12 weeks, respectively. Before November 1, 2022, relative effectiveness against death was 70.3% (95% CI, 32.2 to 87.0), 58.3% (95% CI, 28.5 to 75.7), 41.6% (95% CI, 8.4 to 62.7) and 18.1% (95% CI, 0.0 to 56.9) after 4 weeks, 8 weeks, 12 weeks, and 16 weeks, respectively. After November 1, 2022, relative effectiveness against death was 73.8% (95% CI, 0.0 to 94.8), 52.3% (95% CI, 0.0 to 81.7) and 13.1% (95% CI, 0.0 to 84.3) after 4 weeks, 8 weeks, and 12 weeks, respectively.

## Discussion

Both the Moderna and Pfizer bivalent boosters were associated with additional reduction of Omicron infection in persons who had previously been vaccinated or boosted. Although the two bivalent vaccines targeted the BA.4/BA.5 strains, they were also found to similarly reduce the risks of infection, hospitalization, and death with the BQ.1/BQ.1.1 and XBB/XBB.1.5 strains. The effectiveness against COVID-19 related hospitalization and death was higher than against infection, and it waned gradually over time.

In this study, we evaluated the additional protection of a bivalent booster dose compared with one fewer vaccine dose (i.e., first booster vs. primary vaccination, second booster vs. first booster, or third booster vs. second booster). The effectiveness of bivalent boosters compared with the unvaccinated would be much greater.

A similar study was conducted using the line-level data from the state of North Carolina.^5^ The fundamental conclusions are similar between the two studies. This study covered the time period of September 1, 2022 to June 15, 2023, whereas the North Carolina study covered the time period of September 1, 2022 to February 10, 2023. Thus, this study provided more extensive coverage of the XBB/XBB.1.5 strains.

As with other observational studies, our study is limited by unmeasured confounding bias. In particular, our data did not contain information on underlying medical conditions which might have affected propensity to receive bivalent boosters as well as the risks of infection, hospitalization and death. In addition, we were unable to study the effectiveness of bivalent boosters beyond 9.6 months because the data collection was limited after the end of the federal declaration of emergency.

## Data Availability

Data available upon request from the Nebraska Department of Health and Human Services

